# T1D GRS: a tool for identifying children and adolescents with monogenic diabetes

**DOI:** 10.1101/2025.02.07.25320954

**Authors:** Zuzana Dobiasova, Martina Skopkova, Miloslav Karhanek, Filip Gregus, Miroslav Sabo, Miroslava Huckova, Denisa Lobotkova, Kristina Podolakova, Emilia Jancova, Lubomir Barak, Daniela Gasperikova, Juraj Stanik

**Affiliations:** Diabgene Laboratory, Institute of Experimental Endocrinology, Biomedical Research Center SAS, Dubravska cesta 9, 845 05 Bratislava, Slovakia; Department of Paediatrics, Medical Faculty of the Comenius University and National Institute of Children’s Diseases, Limbova 1, 833 40 Bratislava, Slovakia

**Keywords:** genetic testing, genetic risk score, monogenic diabetes, type 1 diabetes, prioritisation

## Abstract

**The aim** of this study was to determine the effectiveness of the Type 1 Diabetes Genetic Risk Score (T1D GRS) for the prioritisation of children with newly diagnosed hyperglycaemia for genetic testing of monogenic diabetes.

**Methods:** A cohort of 808 children and adolescents with newly diagnosed hyperglycaemia were collected. All underwent standard clinical follow-up and genetic testing based on the knowledge and means accessible at the time. In this cohort and in 189 control subjects (165 monogenic diabetes patients and 24 healthy individuals), we assessed the T1D GRS2 and 10 SNP GRS scores. We assessed the T1D GRS2 cut-off in our cohort and investigated its utility in addition to negative autoantibody status for the prioritisation of cases for genetic testing for monogenic diabetes. Genetic testing included Sanger sequencing, panel sequencing and MLPA.

**Results:** Applying T1D GRS2 in addition to negative autoantibodies on the newly diagnosed hyperglycaemia cohort substantially decreased the number of unnecessarily tested cases. The pick-up rate was increased three-fold, while the sensitivity of the prioritisation decreased only slightly from 77.8% to 72.2% when compared with autoantibodies alone. The majority of monogenic diabetes cases that escaped this prioritisation for genetic testing had low levels of a single autoantibody and were most probably false positives in the autoantibody testing. The monogenic cases that would not be prioritised using GRS2 and autoantibodies were diagnosed based on clinical phenotype. On the other hand, two monogenic diabetes cases with HNF1B-MODY were not originally diagnosed and were identified only thanks to their low GRS2 value.

**Conclusions:** Using T1D GRS in combination with autoantibody testing is effective in decreasing the number of unnecessarily genetically tested cases. This approach, used in addition to the standard clinical evaluation, can be a valuable tool in the early selection of suitable candidates for molecular testing for monogenic diabetes.

**Research in Context:**

- **What is already known about this subject?**
  - Diagnosing monogenic diabetes is important, because gene-tailored treatment is available. In children and adolescents, monogenic diabetes has to be differentiated mostly from type 1 diabetes.
  - Individuals with type 1 diabetes and monogenic diabetes have a different genetic risk for type 1 diabetes.
  - The genetic risk score for type 1 diabetes (T1D GRS) can be computed from disease-associated polymorphisms.
- **What is the key question?**
  - What is the utility of T1D GRS as a tool for the prioritisation of cases for genetic testing of monogenic diabetes?
- **What are the new findings?**
  - More than half of the children with diabetes with negative autoantibodies (type 2 diabetes excluded) and T1D GRS2 below a cut-off was confirmed genetically as having monogenic diabetes.
  - The combination of T1D GRS2 + negative autoantibodies increased the pick-up rate by genetic testing three-fold compared with negative autoantibodies alone, with only small decrease in sensitivity of the prioritisation.
  - Low T1D GRS can draw interest to cases that would otherwise not be suspected of having monogenic diabetes.
- **How might this affect clinical practice in the foreseeable future?**
  - Children and adolescents with confirmed diabetes and normal BMI, negative autoantibodies and low T1D GRS can be prioritised for genetic testing soon after diabetes diagnosis.

## Introduction

The most common forms of diabetes mellitus in children include type 1, type 2 and monogenic diabetes mellitus, mainly represented by MODY (Maturity Onset Diabetes of the Young) [1]. Correct diagnosis of the respective diabetes type is important, as it can influence patient management. In the case of some forms of monogenic diabetes, precision medicine-based treatment is available. Specifically, children with permanent neonatal diabetes mellitus due to *ABCC8* or *KCNJ11* pathogenic variants have a better outcome when insulin is switched to sulfonylurea [2, 3]. Individuals with the *HNF1A* and *HNF4A*-MODY subtypes can also be successfully treated with sulfonylurea, and the *GCK*-MODY type does not require any treatment at all [4, 5]. This explains the need for the active search for monogenic diabetes cases. Distinguishing monogenic diabetes mellitus from other types of diabetes, however, can be challenging. Different approaches are necessary for children compared to the adult population. In adults, monogenic diabetes has to be differentiated mainly from type 2 diabetes and LADA [6]. In children and adolescents, the differential diagnosis from type 1 diabetes, the most common form of diabetes in minors in all European countries [7], is the most challenging.

The prevalence of monogenic diabetes is estimated as 1.1–6.5 % of the paediatric diabetes population [8]. The typical features of monogenic diabetes (early onset, normal BMI, family history of diabetes, measurable C-peptide, and negative pancreatic autoantibodies) are not very helpful in differentiating monogenic diabetes from type 1 diabetes in children and adolescents. Monogenic diabetes and type 1 diabetes share similar age of onset and BMI. The positive family history typical for monogenic diseases is not always present due to *de novo* arising variants [9] and, on the other hand, it is sometimes present in type 1 diabetes due to the important genetic background of this condition [10]. Detectable C-peptide is useful for differentiation only after several years of diabetes duration. Moreover, the low incidence of monogenic diabetes contributes to its misclassification as type 1 diabetes in children. The absence of autoantibodies, which are typically present in type 1 diabetes, is the most useful biomarker in the search of monogenic cases at the time of diabetes onset. However, more than 5 % of children with type 1 diabetes do not have these antibodies [11].

Finally, even when using all available biomarkers and clinical features, monogenic diabetes has to be confirmed by genetic analysis. Due to the high number of antibody-negative type 1 diabetes cases, routine genetic testing based on the absence of autoantibodies is not cost-effective. Patients are thus picked-up using clinical criteria [12] and the expert judgment of experienced clinicians as these criteria have the above-mentioned caveats. Therefore, effort is made to provide additional tools beyond autoantibodies testing that would both reduce the number of cases for genetic testing (increase the positive predictive value) and increase the success rate of diagnosing monogenic diabetes (increase the sensitivity).

A promising possibility for differentiating monogenic and type 1 diabetes emerging in recent years is based on the specific genetic background of type 1 diabetes, which mainly involves specific HLA haplotypes. Individuals with non-autoimmune monogenic forms of diabetes have this genetic background alike the general population and can thus be differentiated from type 1 diabetes cases based on their low genetic risk for this disorder.

The genetic risk can be calculated as a genetic risk score (GRS) which is generally based on single nucleotide polymorphisms (SNPs) associated with given phenotypes in genome-wide association studies (GWAS). Oram et al. proposed a formula for a genetic risk score for type 1 diabetes, T1D GRS1, that included 30 SNPs and discriminated well type 1 and type 2 diabetes [13] or monogenic forms [14]. This was later modified to T1D GRS2 obtained from 67 SNPs with greater discriminatory power [6]. The top ten most weighted SNPs were also proved to provide reasonably high discrimination with lower financial demands, making them more usable in clinical practice [6, 15, 16]

In this paper, we would like to explore the potential of T1D GRS for prioritisation of individuals for genetic testing in addition to autoantibodies.

## Methods

### Study source population

We used three basic cohorts – children with newly diagnosed hyperglycaemia and two control groups – individuals already diagnosed with monogenic diabetes and healthy adults.

#### Children with newly diagnosed hyperglycaemia

This cohort included 808 children of Caucasian origin with newly diagnosed hyperglycaemia who were referred to the Slovak Children Diabetes Centre at the Department of Paediatrics, Faculty of Medicine Comenius University and the National Institute of Children’s Diseases in Bratislava, Slovakia, from 2008 to 2022. The children underwent a standard diagnostic process consisting of measurements of weight, height, BMI, and BMI-SDS along with an evaluation of glycemia monitoring and laboratory findings of HbA1c, C-peptide, pancreatic autoantibodies (GADA, IA-2A, and IAA) and a lipid profile.

#### Control Groups

The control group of individuals already diagnosed with monogenic diabetes (associated with the *GCK, HNF1A, HNF4A, HNF1B, INS, ABCC8, KCNJ11, RFX6, INSR, WFS1*, and *EIF2S3* genes) consisted of 165 subjects and included both children and adults who were referred from outpatient clinics all over Slovakia. Their values for glucose, autoantibodies, HbA1c and serum C-peptide levels measured in local laboratories were taken from questionnaires.

The second control group included 24 healthy individuals over the age of 40 without any history of hyperglycaemia.

### Phenotypes in children with newly diagnosed hyperglycaemia

#### Biochemical analyses

Concentrations of C-peptide, HbA1c and autoantibodies were measured within three days after the patients’ admission to the hospital. C-peptide levels were evaluated using the electrochemiluminescence immunoassay method; HbA1c levels were evaluated from whole blood using the HPLC method, and pancreatic autoantibodies GADA, IA-2A and IAA were detected using ELISA kits [17].

#### ZnT8 autoantibody testing

ZnT8 autoantibodies were tested in patients negative for autoantibodies GADA, anti-IAA and IA-2A, where a plasma sample at the disease onset was available (a subset of 28 children with newly diagnosed hyperglycaemia since the year 2018). The ZnT8 ELISA kit (DRG) was used, and values >15 U/ml were considered positive according to the manufacturer’s instructions.

#### Clinical follow-up

Clinical diagnosis in children with newly diagnosed hyperglycaemia was done at least six months after the initial referral. Diagnosis of type 1 diabetes was based on the current ISPAD criteria [1]. All individuals labelled as type 1 diabetes required ongoing insulin therapy and had at least one of the following features: diabetic ketoacidosis (DKA) at the onset of diabetes, detection of one or more T1D-associated antibodies (GADA, IA-2A, IAA) or an apparent decline of C-peptide serum levels (<200 pmol/l) during the first three years after the diabetes diagnosis. Individuals with central obesity, metabolic syndrome, and without type 1 diabetes-associated antibodies were labelled as type 2 diabetes. Participants with diabetes mellitus but not fulfilling the above-mentioned criteria were labelled as Unclassified. Children with stress hyperglycaemia, or physiological findings were considered Non-diabetic.

### Genotyping for GRS

#### SNP microarray

The DNA extracted from peripheral blood was used for genotyping using the Illumina Infinium Global Screening beadchip GSA MD v3 (Infinium iSelect 24×1 HTS Custom Beadchip Kit), which was performed by the HuGe-F Erasmus University Medical Center, Rotterdam, the Netherlands, as a service.

#### Competitive Allele Specific Polymerase Chain Reaction (KASP) assay

The top ten SNPs with the highest weight that failed to be genotyped by microarray in sufficient quality (with a call score < 0.27) were assessed individually using KASP assay (LGC, Biosearch Technologies). The mutation-specific primers were designed and validated by LGC Genomics (Herts, UK).

### Genetic risk calculation

#### 10 SNP GRS

The 10 SNP GRS was counted as the number of risk-increasing alleles (0, 1, or 2) at the given SNP multiplied by the weight (natural logarithm of the odds ratio) of the SNP as described previously [13].

#### T1D GRS2

The T1D GRS2 was calculated using genotypes for 67 type 1 diabetes-associated SNPs [6]. Variants not included in the GSA array were imputed using the TOPMed (version r2) panel [18], with an RM^2^ threshold of greater than 0.97. Eight SNPs (rs2476601, rs1281934, rs9273342, rs9271346, rs16822632, rs116522341, rs559242105, and rs371250843) were not found in the TOPMed database; therefore, suitable proxy SNPs (rs6679677, rs1281943, rs9273032, rs9271347, rs17840116, rs9268500, rs3129197, and rs9266268) were used instead [19]. The HLA component of GRS2 was calculated using the PRS Toolkit for HLA (version 0.22a) developed by Sharp et al. [6]. The non-HLA component of GRS2 was calculated by taking a weighted sum using odds ratios from Sharp et al. [6], which was then added to the HLA component.

### Genetic analysis of monogenic diabetes genes

All children with unclassified diabetes (n=24) and with clinically defined type 1 diabetes with negative autoantibodies (AAb–) (n=46) underwent genetic testing.

#### Sanger Sequencing

Sequencing of individual genes was done in children with mild fasting hyperglycaemia (*GCK*), in one case with diabetes onset in three months (*ABCC8, KCNJ11* and *INS* genes), and in antibody negative cases with high T1D GRS2 (*HNF1A, INS*).

#### MLPA

In all cases, we performed MLPA (Multiplex Ligation-dependent Probe Amplification) using the SALSA MLPA P241 MODY kit for the detection of large deletions or insertions.

#### Panel sequencing

Panel sequencing was done in all AAb– cases where ZnT8 autoantibodies had been tested and where ZnT8 measurements were unavailable, only in AAb– cases with T1D GRS2 below the cut-off of 11.818. Analysis of 69 genes associated with various forms of monogenic diabetes was done using a custom targeted panel designed by Illumina BaseSpace DesignStudio. Target sequences were exons with 30 bp padding and promoters 300 bp upstream the first exon. The DNA libraries were prepared with Illumina DNA prep with the Enrichment kit and run on iSeq (Illumina). The reads were aligned to the hg19 reference sequence and variants were annotated using the Gemini database [20]. Coverage of the main monogenic diabetes genes (*HNF1A, HNF1B, HNF4A, GCK, INS, NEURO1, PDX1, ABCC8, KCNJ11, CEL, RFX6, mt*.*3243A>G*) was checked and individual exons were sequenced by Sanger sequencing in the case of < 6 reads.

#### Variant classification

We classified the identified variants according to the gene-specific ClinGen Monogenic Diabetes Expert Panel Specifications to the ACMG/AMP Variant Interpretation Guidelines for genes *GCK* (version 1.3.0), *HNF1A* (version 2.1.0.) and *HNF4A* (version 2.0.0) (https://cspec.genome.network/cspec/ui/svi/affiliation/50016/). Variants in other considered genes were classified using ACMG guidelines [21], with modifications as recommended by the ClinGen SVI committee (https://clinicalgenome.org/working-groups/sequence-variant-interpretation/).

### Statistics

Metric data were checked for normality using the Shapiro–Wilk test. Normally distributed data are expressed as the mean±SD. Non-normally distributed data (i.e., BMI-SDS and C-peptide) are expressed as the median and interquartile range. Non-normally distributed data were logarithmically transformed prior to further statistical analyses. Confidence intervals for percentages in binary data were calculated using the Wilson/Brown method. Differences between groups in Table 1 and Figure 2 were tested using ANOVA with the Tukey test for *post-hoc* analyses for metric data and the chi-square test with Fisher’s test in *post-hoc* analyses for binary data.

**Table 1.** Characteristics of the basic cohorts (A), of the groups created from the newly diagnosed hyperglycaemic children after clinical follow-up and genetic testing (B), and T1D individuals with positive (AAb+), negative (AAb-) or unknown (AAbNA) autoantibodies. In the *post hoc* analyses, groups were compared to the first group listed - i.e. to “Hyperglycaemia” in A, to “T1D” in B, and to AAb+ in C. ***p<0.001, **p<0.01, *p<0.05

|  | n | GRS2<br>[mean ±SD] | 10 SNP GRS<br>[mean ±SD] | male<br>[%] (95CI) | Age at<br>onset<br>[years ±SD] | Auto-<br>antibodies<br>[%] (95CI) | BMI-SDS at onset<br>[kg/m2] (IQR)<sup>a</sup> | HbA1c<br>at onset<br>[%] | HbA1c<br>at onset<br>[mmol/mol±SD] | C-peptide at<br>onset [pmol/l]<br>(IQR)<sup>a</sup> | DKA<br>[%] (95CI) | Treatment<br>with insulin<br>[%] (95CI) |
| --- | --- | --- | --- | --- | --- | --- | --- | --- | --- | --- | --- | --- |
| **A** |  |  |  |  |  |  |  |  |  |  |  |  |
| **Basic cohorts** |  |  |  |  |  |  |  |  |  |  |  |  |
| Hyperglycaemia | 808 | 13.3±2.09 | 11.9±1.6 | 55.2<br>(51.7-58.6) | 9.41±4.77 | 87.6<br>(85.0-89.8) | -0.418<br>(-1.045;0.357) | 11.4±1.7<br>6 | 101.0±30.2 | 163.0<br>(94.0;266.0) | 35.4<br>(32.1-38.9) | 97.3<br>(95.9-98.3) |
| Monogenic controls | 165 | **9.72±2.16\*\*\*** | **9.53±1.88\*\*\*** | 49.7<br>(42.2-57.2) | **11.7±6.32\*\***<br>* | **0.9**<br>**(0.04-4.8)\*\*\*** | **0.161**<br>**(-0.543;0.900)\*\*\*** | 6.79±1.6<br>2 | **50.7±17.8\*\*\*** | **433.0**<br>**(285.5;656.0)\*\*\*** | **0.8**<br>**(0.0-4.3)\*\*\*** | N/A |
| Healthy controls | 24 | **9.71±2.49\*\*\*** | **9.21±2.06\*\*\*** | 50.0<br>(31.4-68.6) | N/A | N/A | **1.043**<br>**(-0.154;2.536)\*\*\*** | N/A | N/A | N/A | NA | N/A |
| **B** |  |  |  |  |  |  |  |  |  |  |  |  |
| **Hyperglycaemia group**<br>**after follow-up and**<br>**genetics** |  |  |  |  |  |  |  |  |  |  |  |  |
| T1D | 715 | 13.7±1.72 | 12.1±1.43 | 55.7<br>(52.0-59.3) | 9.34±4.5 | 93.6<br>(91.5-95.2) | -0.456<br>(-1.066;0.299) | 11.7±2.5 | 105.0±37.3 | 152.5<br>(88.3;238.0) | 36.7<br>(33.2-40.4) | 100.0<br>(99.5-100.0) |
| T2D | 13 | **9.48±2.09\*\*\*** | **9.7±1.24\*\*\*** | 61.5<br>(35.5-82.3) | **14.0±2.82\*\*** | **15.4**<br>**(2.7-42.2)\*\*\*** | 5.060<br>(3.329;6.698)*** | 9.96±2.3<br>4 | 85.4±25.5 | **1296.0**<br>**(1006.0;2010.5)\*\*\*** | **0.0**<br>**(0.0-22.8)\*\*** | **30.8**<br>**(12.7-57.6)\*\*\*** |
| New monogenic | 18 | **9.83±1.91\*\*\*** | **9.58±1.45\*\*\*** | 44.4<br>(24.6-66.3) | 9.43±5.62 | **22.2**<br>**(9.0-45.2)\*\*\*** | -0.476<br>(-1.051;-0.119) | 7.32±3.1<br>2 | **56.5±34.1\*\*\*** | **409.0**<br>**(273.0;526.0)\*\*\*** | **5.6**<br>**(0.3-25.8)\*\*** | **37.5**<br>**(18.5-61.4)\*\*\*** |
| MODY-VUS | 2 | 11.3±2.61 | 10.4±0.247 | 100.0<br>(17.8-100) | 11.6±0.59 | **0.0**<br>**(0.0-82.2)\*\*** | 0.963;-1.110 | 14.2±1.3<br>4 | 131.0±14.7 | 227.5;121.0 | 100.0<br>(17.8-100.0) | 100.0<br>(17.8-100.0) |
| Unclassified | 6 | **11.1±1.91\*\*** | **10.1±1.71\*** | 83.3<br>(43.7-99.1) | 13.9±3.82 | **16.7**<br>**(0.9-56.3)\*\*\*** | 1.281<br>(-0.901;2.467) | 7.48±2.4<br>3 | **58.3±26.5\*\*\*** | **666.5**<br>**(408.0;1160.8)\*\*\*** | 16.7<br>(0.9-56.3) | **16.7**<br>**(0.9-56.4)\*\*\*** |
| Non-DM | 19 | **8.87±2.4\*\*\*** | **8.96±1.41\*\*\*** | 52.6<br>(31.7-72.7) | NA | **22.2**<br>**(9.0-45.2)\*\*\*** | -0.035<br>(-0.734;0.577) | 5.19±0.2<br>4 | **33.3±2.58\*\*\*** | **390.5**<br>**(278.0;603.5)\*\*\*** | **0.0**<br>**(0.0-16.8)\*\*\*** | **0.0**<br>**(0.0-16.8)\*\*\*** |
| **C** |  |  |  |  |  |  |  |  |  |  |  |  |
| **T1D group divided**<br>**according to AAb status** |  |  |  |  |  |  |  |  |  |  |  |  |
| AAb+ | 643 | 13.6±1.73 | 12.1±1.42 | 56.0<br>(52.1-59.8) | 9.48±4.48 | 100.0<br>(99.4-100.0) | -0.444<br>(-1.071;0.313) | 11.7±2.5 | 105±27.3 | 157.0<br>(91.0;241.0) | 37.0<br>(33.4-40.8) | 100.0<br>(99.4-100.0) |
| AAb- | 44 | 14.1±1.75 | 12.3±1.72 | 56.8<br>(42.2-70.3) | 8.02±4.2 | **0.0**<br>**(0.0-8.0)\*\*\*** | -0.570<br>(-1.098;0.098) | 11.9±2.2 | 107±24.1 | 123.5<br>(74.0;206.3) | 31.8<br>(20.0-46.6) | 100.0<br>(92.0-100.0) |
| AAbNA | 28 | 13.9±1.4 | 12.3±1.17 | 46.4<br>(29.5-64.2) | 8.2±5.04 | N/A | -0.638<br>(-0.900;-0.004) | 11.5±1.9<br>5 | 102±21.4 | 114.0<br>(87.5;195.0) | 38.9<br>(20.3-61.8) | 100.0<br>(87.9-100.0) |

<sup>a</sup>analyzed as log transformed

For the generation of plots, the *ggplot2* v3.5.1 package for R was used [22]. Boxplots present medians and interquartile ranges. ROC analysis was performed using the *Optimal*.*Cutpoints* v1.1-5 package in R and optimal cut-off was estimated based on the highest Youden index [23, 24].

### Ethics

The study was approved by the Ethics Committee of the National Institute of Children’s Health in Bratislava, Slovakia (EK1152022), and adhered to the tenets outlined in the Declaration of Helsinki, as revised in 2008. Written informed consent was obtained from all subjects and/or their legal guardian(s).

## Results

We aimed to assess the usefulness of T1D GRS for the prioritisation of children with newly diagnosed hyperglycaemia for genetic testing of monogenic diabetes. The goal was to obtain an optimal ratio of sensitivity and specificity that would allow to identify most monogenic cases and to reduce the number of expensive genetic analyses simultaneously. The studied subjects underwent the standard diagnostic pipeline including clinical evaluation and genetic testing in the case of suspicion on monogenic diabetes. The T1D GRS estimate and its use in prioritisation for genetic testing was done retrospectively.

### Clinical follow-up

The children with newly diagnosed hyperglycaemia (n=808) were first assessed clinically (without knowledge of T1D GRS or genetic testing) after at least six months of follow-up and a clinical diagnosis of type 1 (n=717) or type 2 diabetes (n=13) was made. Twenty-four individuals were labelled as Unclassified, and diabetes was not confirmed in 19 cases. Thirty-five cases were lost to follow-up and were thus excluded from the final comparisons (Fig. 1a).

**Fig. 1:**
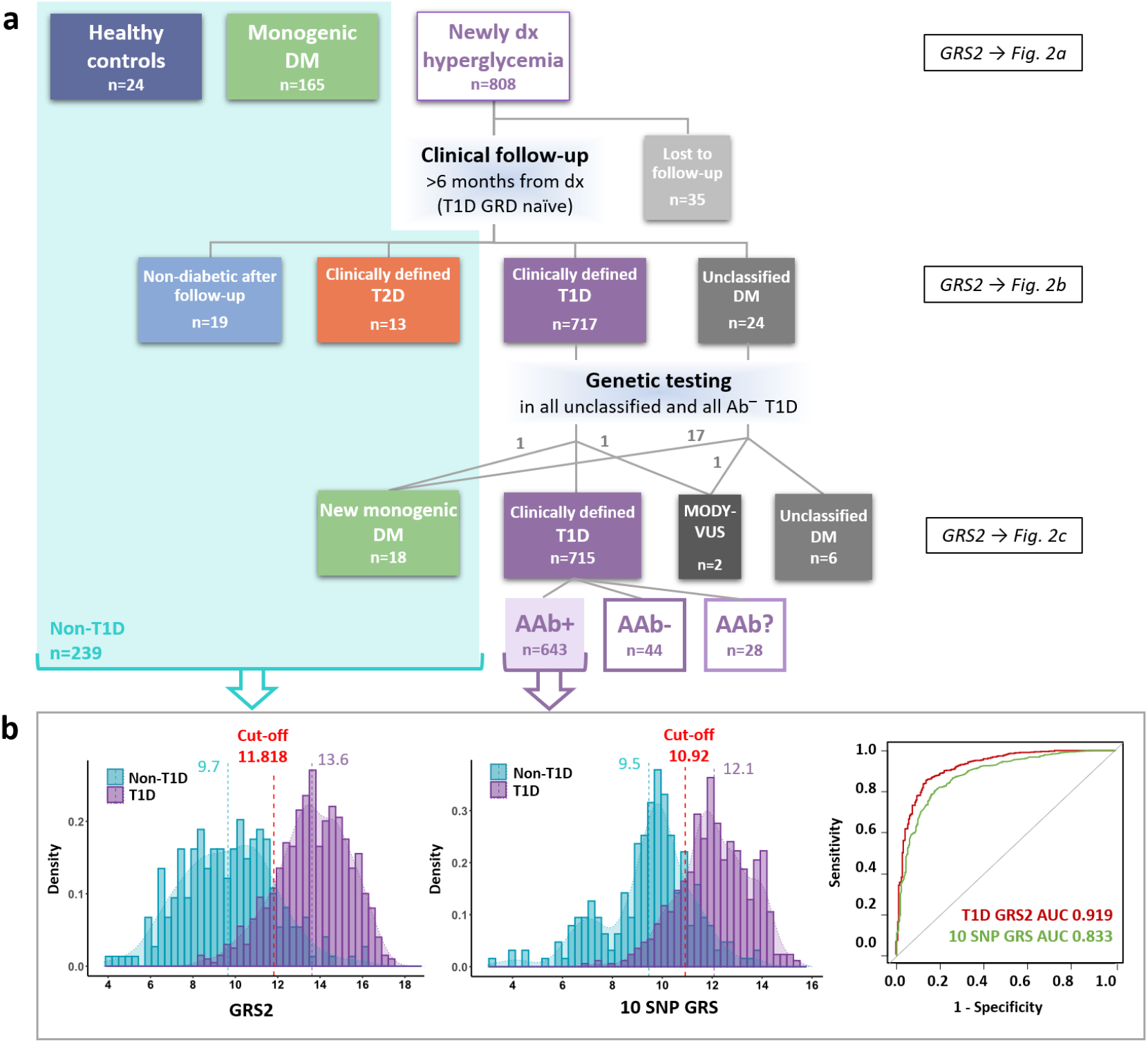
a) Workflow describing the grouping based on clinical follow-up and genetic testing and the selection of individuals for T1D GRS2 cut-off estimation. b) Histograms and ROC curves of T1D GRS2 and 10 SNP GRS values in non-T1D individuals (n=239) versus AAb+ T1D cases (n=643). Mean values of the two groups and the cut-offs are depicted in the histograms.

**Fig. 2:**
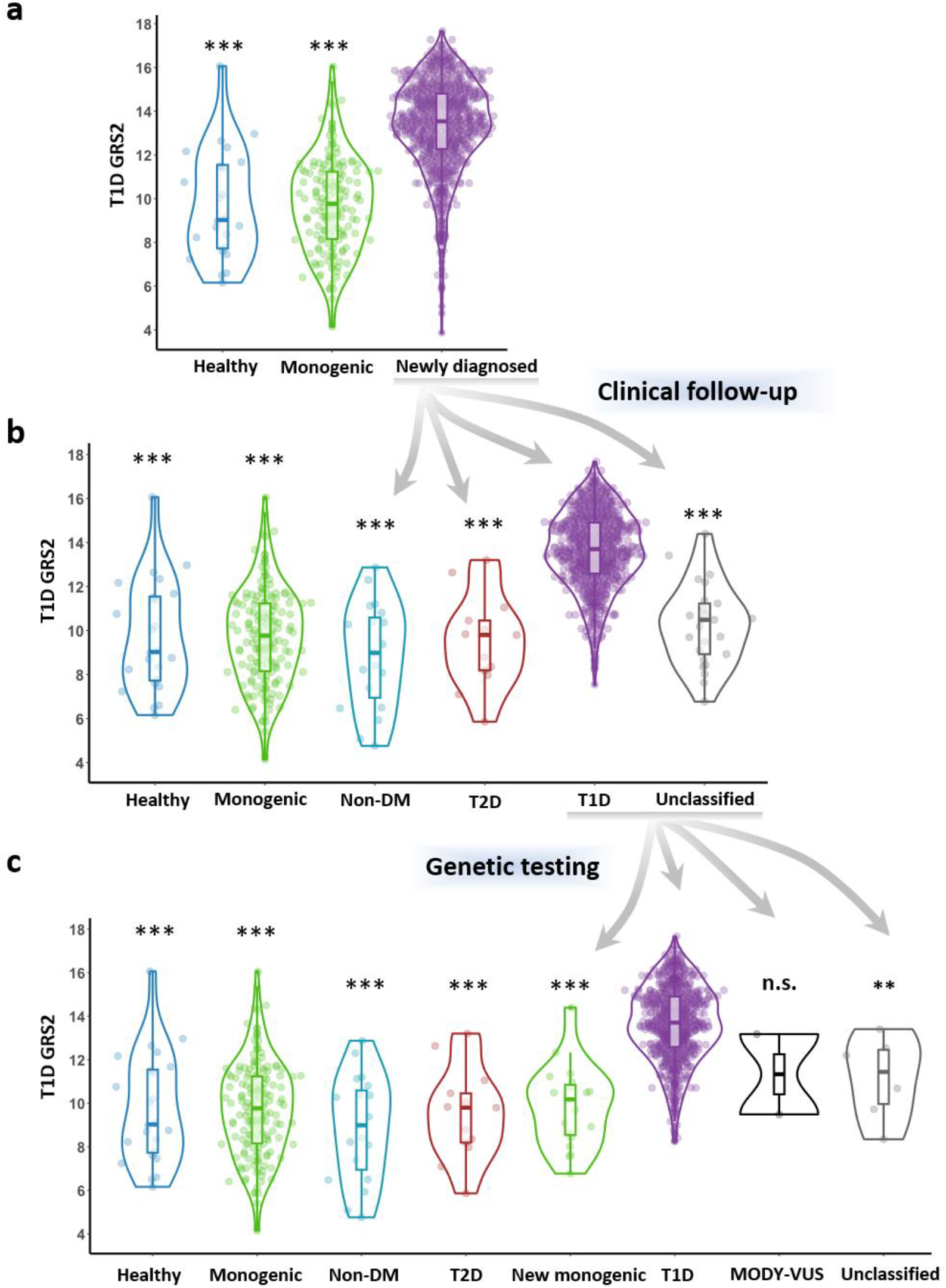
a) Distribution of the T1D GRS2 values in the basic cohorts; b) in the groups after clinical follow-up and c) in the groups formed after genetic testing. ***p<0.001, **p<0.01, n.s. – not significant; ANOVA and post-hoc Tukey (a) vs. newly diagnosed group, (b) and (c) vs. T1D group.

The clinical characteristics of these groups, as well as the control groups, are summarised in Table 1.

### T1D genetic risk score cut-off estimation

T1D GRS2 was assessed in all subjects (n=997) included in the study (Table 1). Figure 2a shows the distribution of T1D GRS within the basic cohorts. As expected, T1D GRS2 of the cohort with monogenic diabetes (9.72±2.16, n=165) was comparable with healthy individuals (9.71±2.49, n=24), while it was significantly higher (13.3±2.09, n=808; p<0.001) in the children with newly diagnosed hyperglycaemia, where the a priori risk of type 1 diabetes is the highest.

After the clinical follow-up (Fig. 2b), the T1D GRS2 in the newly created groups with type 2 diabetes (T2D, 9.48±2.09, n=13) or without confirmed diabetes (Non-DM, 8.87±2.40, n=19) were comparable with healthy individuals and significantly different from the clinically defined T1D group (13.6±1.74, n=717, p<0.001). The T1D GRS2 of the unclassified diabetes group (10.2±1.88, n=24) was significantly different from the T1D groups as well (p<0.001) and was comparable with other non-T1D groups (no significant differences). This indicates a higher possibility of monogenic diabetes cases being found in this group.

After genetic analysis, a group of newly identified individuals with monogenic diabetes was formed whose GRS2 was significantly different from the T1D group (9.83 ±1.91, n=18, p<0.001) (Fig. 2c, Table 1).

All non-T1D groups, except for the children who were lost to follow-up and children with unclassified diabetes, were used for the T1D GRS2 cut-off estimation (n=239) (Fig. 1a). To be sure that no monogenic cases were included, only autoantibody positive (AAb+) individuals were counted in the T1D group (n=643). The ROC analysis showed very good discrimination of type 1 diabetes cases (AUC 0.919), and the optimal cut-off point based on the highest Youden index (0.716) was set at 11.818 (Fig. 1b).

As 10 SNP GRS would be more practical for use in clinical practice, we also computed this score to determine its efficacy. The discrimination of type 1 diabetes and non-type 1 diabetes individuals was lower (AUC 0.883, Youden 0.631) but still reasonably high, with the cut-off set at 10.92 (Fig. 1b).

### Genetic testing

All children with unclassified diabetes (n=24) and with clinically defined type 1 diabetes with negative autoantibodies (AAb–) (n=46) underwent genetic testing (Fig. 1a).

In summary, 18 cases with monogenic diabetes (14 GCK-MODY, 2 HNF1B-MODY, 1 HNF1A-MODY and 1 INS-PND) were confirmed among 70 children with either clinically diagnosed type 1 diabetes without autoantibodies (n=1) or with unclassified diabetes (n=17) (Fig. 1a). The clinical phenotypes of these cases are summarised in Table 2.

**Table 2.**
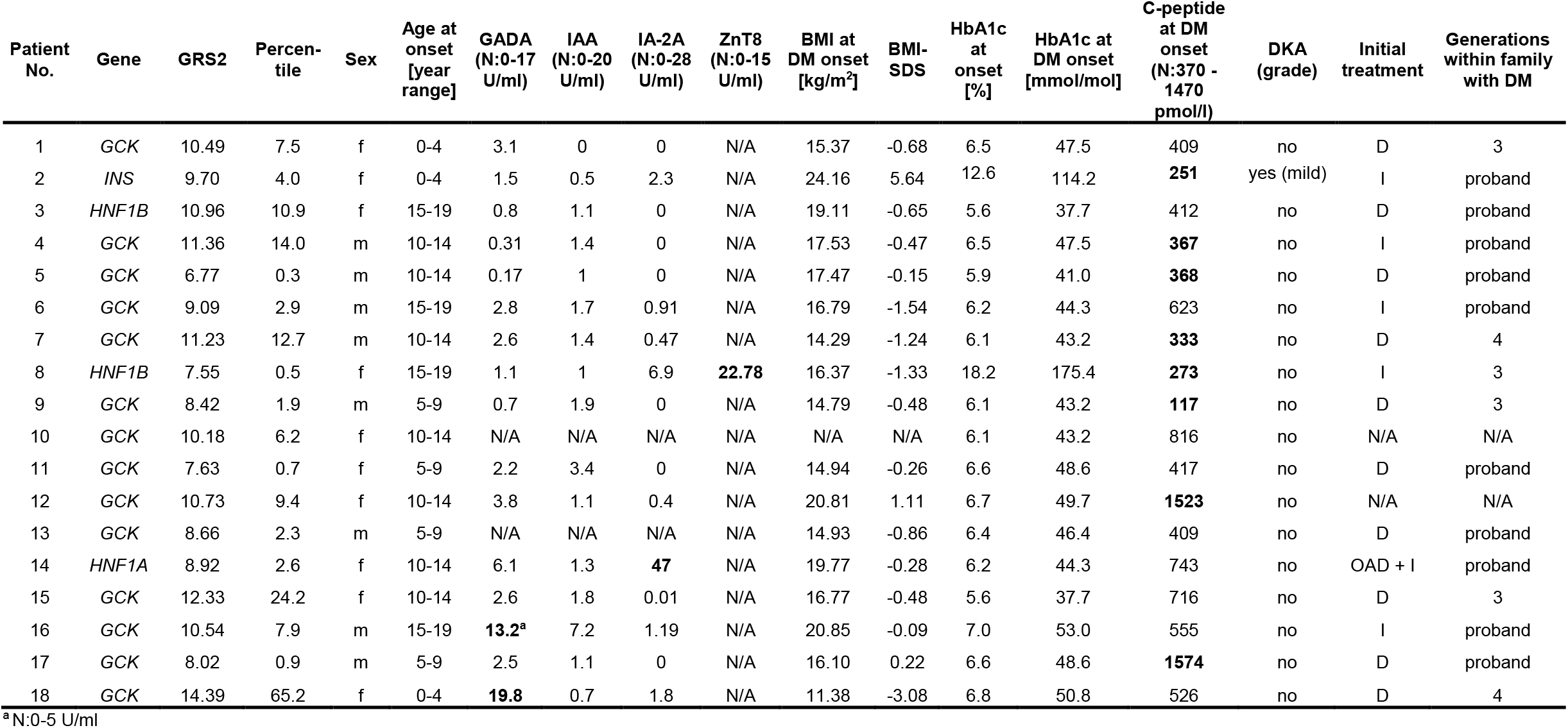
Characteristics of individuals with monogenic diabetes identified among the children with newly diagnosed hyperglycaemia. Percentile is counted from all children with diabetes from the newly diagnosed cohort (n=754) (T2D included). Values in bold are outside the normal range. *Abbreviations: D – diet, DKA – diabetic ketoacidosis, DM – diabetes, I – insulin, N – normal range, N/A – not available, OAD – oral antidiabetics*

| Patient No. | Gene | GRS2 | Percentile | Sex | Age at onset [year range] | GADA (N:0-17 U/ml) | IAA (N:0-20 U/ml) | IA-2A (N:0-28 U/ml) | ZnT8 (N:0-15 U/ml) | BMI at DM onset [kg/m<sup>2</sup>] | BMI-SDS | HbA1c at onset [%] | HbA1c at DM onset [mmol/mol] | C-peptide at DM onset (N:370 - 1470 pmol/l) | DKA (grade) | Initial treatment | Generations within family with DM |
| --- | --- | --- | --- | --- | --- | --- | --- | --- | --- | --- | --- | --- | --- | --- | --- | --- | --- |
| 1 | *GCK* | 10.49 | 7.5 | f | 0-4 | 3.1 | 0 | 0 | N/A | 15.37 | -0.68 | 6.5 | 47.5 | 409 | no | D | 3 |
| 2 | *INS* | 9.70 | 4.0 | f | 0-4 | 1.5 | 0.5 | 2.3 | N/A | 24.16 | 5.64 | 12.6 | 114.2 | **251** | yes (mild) | I | proband |
| 3 | *HNF1B* | 10.96 | 10.9 | f | 15-19 | 0.8 | 1.1 | 0 | N/A | 19.11 | -0.65 | 5.6 | 37.7 | 412 | no | D | proband |
| 4 | *GCK* | 11.36 | 14.0 | m | 10-14 | 0.31 | 1.4 | 0 | N/A | 17.53 | -0.47 | 6.5 | 47.5 | **367** | no | I | proband |
| 5 | *GCK* | 6.77 | 0.3 | m | 10-14 | 0.17 | 1 | 0 | N/A | 17.47 | -0.15 | 5.9 | 41.0 | **368** | no | D | proband |
| 6 | *GCK* | 9.09 | 2.9 | m | 15-19 | 2.8 | 1.7 | 0.91 | N/A | 16.79 | -1.54 | 6.2 | 44.3 | 623 | no | I | proband |
| 7 | *GCK* | 11.23 | 12.7 | m | 10-14 | 2.6 | 1.4 | 0.47 | N/A | 14.29 | -1.24 | 6.1 | 43.2 | **333** | no | D | 4 |
| 8 | *HNF1B* | 7.55 | 0.5 | f | 15-19 | 1.1 | 1 | 6.9 | **22.78** | 16.37 | -1.33 | 18.2 | 175.4 | **273** | no | I | 3 |
| 9 | *GCK* | 8.42 | 1.9 | m | 5-9 | 0.7 | 1.9 | 0 | N/A | 14.79 | -0.48 | 6.1 | 43.2 | **117** | no | D | 3 |
| 10 | *GCK* | 10.18 | 6.2 | f | 10-14 | N/A | N/A | N/A | N/A | N/A | N/A | 6.1 | 43.2 | 816 | no | N/A | N/A |
| 11 | *GCK* | 7.63 | 0.7 | f | 5-9 | 2.2 | 3.4 | 0 | N/A | 14.94 | -0.26 | 6.6 | 48.6 | 417 | no | D | proband |
| 12 | *GCK* | 10.73 | 9.4 | f | 10-14 | 3.8 | 1.1 | 0.4 | N/A | 20.81 | 1.11 | 6.7 | 49.7 | **1523** | no | N/A | N/A |
| 13 | *GCK* | 8.66 | 2.3 | m | 5-9 | N/A | N/A | N/A | N/A | 14.93 | -0.86 | 6.4 | 46.4 | 409 | no | D | proband |
| 14 | *HNF1A* | 8.92 | 2.6 | f | 10-14 | 6.1 | 1.3 | **47** | N/A | 19.77 | -0.28 | 6.2 | 44.3 | 743 | no | OAD + I | proband |
| 15 | *GCK* | 12.33 | 24.2 | f | 10-14 | 2.6 | 1.8 | 0.01 | N/A | 16.77 | -0.48 | 5.6 | 37.7 | 716 | no | D | 3 |
| 16 | *GCK* | 10.54 | 7.9 | m | 15-19 | **13.2<sup>a</sup>** | 7.2 | 1.19 | N/A | 20.85 | -0.09 | 7.0 | 53.0 | 555 | no | I | proband |
| 17 | *GCK* | 8.02 | 0.9 | m | 5-9 | 2.5 | 1.1 | 0 | N/A | 16.10 | 0.22 | 6.6 | 48.6 | **1574** | no | D | proband |
| 18 | *GCK* | 14.39 | 65.2 | f | 0-4 | **19.8** | 0.7 | 1.8 | N/A | 11.38 | -3.08 | 6.8 | 50.8 | 526 | no | D | 4 |

<sup>a</sup> N:0-5 U/ml

### Evaluation of T1D GRS2’s usefulness for prioritisation for genetic testing

To evaluate the use of T1D GRS for prioritisation, we took only subjects from the newly diagnosed hyperglycaemia cohort from the Slovak Children Diabetes Centre who were evaluated as type 1 diabetes, unclassified or as monogenic DM after genetic testing, where antibody status was known (n=711, 18 monogenic, 6 unclassified, and 687 type 1 diabetes cases). Two MODY-VUS cases were not included due to uncertainty. A summary of the outcomes of prioritisation is shown in Table 3.

**Table 3.** Outcomes of the prioritisation of cases for genetic testing based on autoantibodies alone, T1D GRS2 alone, and their combination, as well as based on 10 SNP GRS alone and in combination with autoantibodies. Individuals with known autoantibody status from the newly identified hyperglycaemia groups were included (T2D cases and MODY-VUS excluded) (n=711). Subjects fulfilling the tested criterion were considered positive, i.e., prioritised for genetic testing. True positives were monogenic cases that would be prioritised and false positives were cases that would be prioritised for monogenic testing but without any genetic cause confirmed. False negatives were monogenic cases that would not be prioritised and remaining were true negatives. *Abbreviations: AAb – autoantibodies, AI – accuracy index, DLR-N – negative diagnostic likelihood ratio, DLR-P – positive diagnostic likelihood ratio, FP – false positive, FN – false negative, GRS2 – T1D genetic risk score 2, N – number, NPV – negative predictive value, PPV – positive predictive value, Se – sensitivity, Sp – specificity, TP – true positive*.

| *Criterion* | *N of prioritized from n=711* | *N of TP (identified monogenic)* | *N of FP (tested unnecessarily)* | *N of FN (mistakenly not tested)* | *Se* | *Sp* | *PPV* | *NPV* | *DLR-P* | *DLR-N* | *AI* |
| --- | --- | --- | --- | --- | --- | --- | --- | --- | --- | --- | --- |
| **AAb–** | 63 | 14 | 49 | 4 | 0.778 | 0.929 | 0.222 | 0.994 | 11.0 | 0.239 | 0.925 |
| **GRS2 below cut-off** | 119 | 16 | 103 | 2 | 0.889 | 0.851 | 0.134 | 0.997 | 5.98 | 0.131 | 0.852 |
| **GRS2 + AAb–** | 21 | 13 | 8 | 5 | 0.722 | 0.988 | 0.619 | 0.993 | 62.56 | 0.281 | 0.982 |
| **10 SNP GRS below cut-off** | 152 | 16 | 136 | 2 | 0.889 | 0.804 | 0.105 | 0.996 | 4.53 | 0.138 | 0.806 |
| **10 SNP GRS + AAb–** | 25 | 13 | 12 | 5 | 0.722 | 0.983 | 0.52 | 0.993 | 41.71 | 0.283 | 0.976 |

First, we compared the use of T1D GRS2 (tested if <11.818) with that of autoantibodies (tested if AAb–), each individually (Table 3). The sensitivity of using autoantibodies alone was lower than using T1D GRS2 alone due to the presence of four false negative cases. Upon retrospective investigation of these cases, it was shown that all of them had been tested positive with only low levels of a single autoantibody (2x GADA, 1x IA-2A, 1x ZnT8 – details in Table 2). Other test characteristics, such as the specificity, accuracy index, positive predictive value and positive diagnostic likelihood ratio, were higher using autoantibodies than GRS2 alone (Table 3). Still, 103 of 119 subjects with T1D GRS2 below the cut-off or 49 of 63 newly diagnosed AAb– cases would have been genetically tested unnecessarily.

Subsequently, we simulated prioritisation for genetic testing based on both negative autoantibodies and T1D GRS2 below the cut-off (Fig. 3a, Table 3). Of 63 AAb– cases, 21 had T1D GRS2 under the cut-off and thus fulfilled the prioritisation criteria. Only eight of them were false positives from the T1D or unclassified group (positive predictive value 0.636) and 13 of 18 monogenic cases would be tested using this prioritisation (sensitivity 72.2%). The five false negative cases consisted of those four individuals with low levels of a single antibody mentioned above, and one of them also had a GRS2 above the cut-off; plus, one case was AAb– but with GRS2 above the cut-off. Using this strategy, the prioritisation had the highest specificity (98.8%), accuracy index (0.982) and a positive diagnostic likelihood ratio of 62.56.

**Fig. 3.**
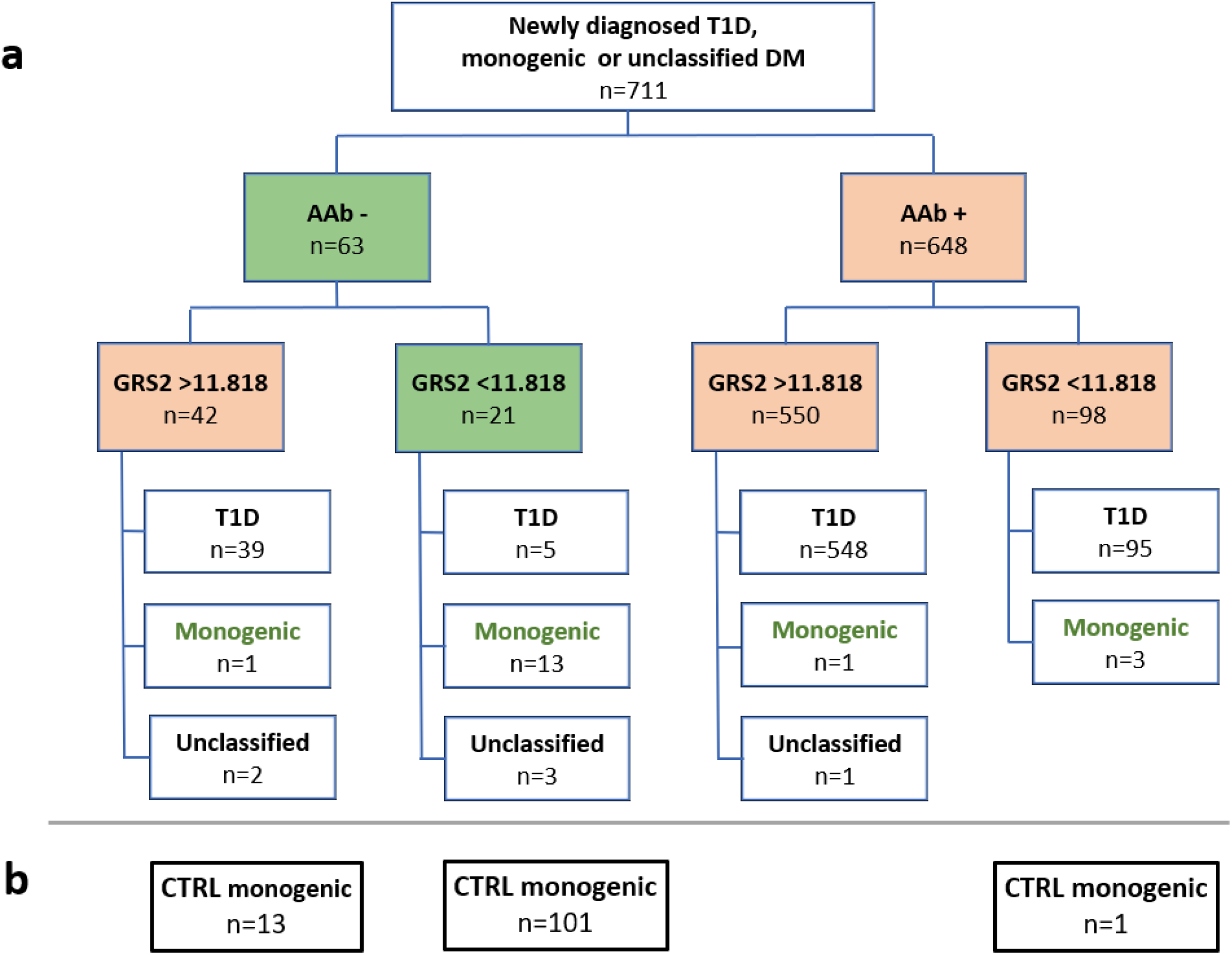
a) Stratification of newly identified children with diabetes (T2D excluded) according to autoantibodies and T1D GRS2; b) Distribution of the monogenic diabetes cases from the control cohort according to autoantibodies and T1D GRS2.

The positive diagnostic likelihood ratio remained high (74.25) even when we added 115 already identified control monogenic cases with known autoantibody status to the simulation (Fig. 3b). Of these 115 cases, 101 would be tested by this approach (sensitivity 87.8%). The 14 remaining control monogenic cases presented false negatives. One monogenic case would not have been tested due to positive autoantibodies. This was shown to be a child with *RFX6* biallelic monogenic diabetes who had positive IAA antibodies detected five years after initiation of insulin therapy. The remaining false negatives were 13 control monogenic cases with T1D GRS2 above the 11.818 cut-off point.

When we simulated the same prioritisation process with the newly diagnosed hyperglycaemia cohort using 10 SNP GRS, which would be more readily available for clinical practice then the T1D GRS2, the number of false negatives and sensitivity remained the same as using the GRS2 computed from 67 SNPs (Supplementary Figure 1a, Table 3). The number of unnecessarily tested individuals increased from 8 to 12, but that is still much lower than 49 false positive individuals prioritised through autoantibodies alone. Of the control monogenic diabetes cases, only 91 would be tested (sensitivity 79.1%), 23 had 10 SNP GRS above the cut-off, and one case was AAb+ (Supplementary Figure 1b).

## Discussion

In this study, we identified the T1D GRS2 cut-off for the Slovak population and applied it, in addition to AAb status and clinical evaluation, for the prioritisation of children with newly diagnosed hyperglycaemia for genetic testing.

### T1D GRS for differentiation between DM types

Until now, published studies focused on using T1D GRS for differentiation between type 1 diabetes and other types of diabetes in young adults or a mixed population. Oram et al. [13] showed that T1D GRS based on 30 SNPs successfully differentiates type 1 from type 2 diabetes (AUC 0.88) and Patel et al., 2016 [14] showed it can be useful in the differentiation of MODY (AUC 0.87) and neonatal diabetes from type 1 diabetes [14]. Both studies were done on a British or white European population. These results were confirmed on Iranian population using the top nine most informative SNPs [25]. The Iranian study analysed a population of children with high consanguinity, hence another spectrum of monogenic diabetes genetic causes.

In 2019, an improved T1D GRS2 score was published which reached a very high power (AUC 0.927) for discrimination of type 1 diabetes in case and control subjects from the Type 1 Diabetes Genetics Consortium [6], and our results are in line with these results. We have confirmed the high discriminatory power of the T1D GRS2 (AUC 0.919), as well as of the more accessible 10 SNP GRS (AUC 0.883) in our cases and controls from Slovakia.

T1D GRS seems to be population-specific, which corresponds with the fact that different populations have different genetic predisposition for type 1 diabetes. The GRS2 value was shown to be different in African-American and European-American populations [26], and thus the individual values should be interpreted in the context of the appropriate population. The mean GRS2 values in our T1D group (both median and mean 13.7) are somewhat lower than those found in the British population (median 14.6 in [6] or mean 14.9 in [27]. The GRS2 mean value was 9.7 in the non-T1D group (n=239) in our study, which is also lower than 10.3 previously found in the control European ancestry [27]. The difference can be due to slightly different genetic background in Slavic population, but the role of a low number of individuals in our control cohort cannot be excluded.

### Combination of T1D GRS with other parameters

Usually, clinical criteria, such as autoantibodies, BMI, age of onset, C-peptide values, family history [28] or tools such as the MODY calculator [29], are used for decisions on whether or not to test a given patient. However, these parameters have caveats mentioned above, which results in either a lower pick-up rate or missing the diagnosis in atypical cases when applying criteria that are too loose or too strict, respectively. The low T1D GRS in non-T1D cases, as we show, can be successfully used as an additional piece of information for prioritising patients for genetic testing. Sharp et al. [6] showed that a combination of GRS and phenotypic features, such as BMI, antibodies and age of onset, improved the discrimination of type 1 and type 2 diabetes. BMI and age of onset are very similar for monogenic and type 1 diabetes in children and adolescents (Table 1). Therefore, we only combined GRS with autoantibodies in addition to clinical assessment for the differential diagnosis of monogenic diabetes from type 1 diabetes to increase the pick-up rate by genetic testing. This approach would not be necessary for discrimination of clinically well-defined monogenic diabetes types such as GCK-MODY, but it can be more important in the case of clinically more similar types, such as HNF-MODY and type 1 diabetes.

As our data show, the discriminatory power of autoantibodies or GRS2 is already very high individually, but their combination dramatically decreases the number of cases that would be genetically tested unnecessarily (from 103 in AAbs or 49 in GRS2 to 8 if GRS2 and AAbs were combined). Adding T1D GRS2 into consideration for genetic testing of non-T2D AAb– cases increased the pick-up rate substantially – from 22% to 62% of cases (Fig. 3a, Table 3), while the sensitivity decreased only marginally, from 77.8% to 72.2% (Table 3) (four and five false negatives, respectively). Four individuals with monogenic diabetes would not be tested using exclusively this approach, due to most probably false positive autoantibodies testing. The risk of using autoantibodies is in unreliably determined reference ranges. As also shown by our data, the normal-range should be assessed for each method and population and borderline AAb values should be considered cautiously, in the context of other clinical and genetic data. Based on our experience, low T1D GRS could be an impulse for retesting in the case of a single positive autoantibody or initiation of genetic testing in spite of borderline levels. Moreover, in cases with monogenic autoimmune diabetes, autoantibodies are frequently positive, with overlapping autoimmune conditions. The timing of autoantibodies measurement is also important. Autoantibodies are useful for diagnosing children, but their levels may decrease during life and can be less useful for diagnosis several years after DM onset. On the other hand, IAA autoantibodies can rise after insulin therapy [30], which was probably the case in one of our IAA-positive patients from monogenic control group with the *RFX6* bi-allelic pathogenic variants. C-peptide, in contrast, is useful only several years after diabetes onset, when its levels usually decrease in type 1 diabetes, but remain preserved in monogenic diabetes types [7]. There are more biomarkers that are studied with the aim of better monogenic diabetes diagnostics, such as hsCRP [31, 32] or glycosylation patterns [33], but they are useful only for the HNF1A-MODY subtype identification. In contrast to these biomarkers, low T1D GRS is common for all monogenic diabetes subtypes and is constant throughout the life span and can thus be used in novel cases as well as in cases with onset farther in the past.

The sensitivity of using T1D GRS2 and AAb– for the prioritisation of monogenic cases in the new hyperglycaemia cohort (72.2%) was not only lower due to positive autoantibodies but also due to one monogenic case with GRS2 above the threshold (Fig. 3a). Indeed, when we applied this stratification to the control group of monogenic diabetes cases, 13 of 115 cases (11%) had GRS2 above the cut-off. If we would like to increase the sensitivity to include more AAb-monogenic cases in genetic testing, we would have to increase the T1D GRS2 cut-off. The stringency of the cut-off used depends on financial limitations, however, presence of monogenic diabetes does not exclude presence of high T1D GRS2, or even type 1 diabetes, and vice versa. It is therefore not reasonable to increase the cut-off to comprise all the monogenic cases in the tested population. Moreover, a majority of the identified monogenic cases in this study were genetically tested already based on their clinical picture. Only two cases with HNF1B-MODY escaped the standard diagnostic pipeline and were found only thanks to low T1D GRS2. Neither of these two children had known abnormalities of the uropoetic tract, which was confirmed only after the genetic diagnosis.

### Application to the clinical practice

In real-world clinical practice, initial differential diagnostics is done using clinical criteria, where clinically well-defined subtypes, such as GCK-MODY can be identified. Consequently, for clinically unclear cases, which are not as readily differentiated from type 1 diabetes, GRS2 can be a valuable tool for decision-making. The high discriminatory power of T1D GRS2 is great, but in clinical practice the use of 10 SNP GRS is more convenient and accessible, either through individual SNP testing or using biochips [16]. In our cohort, using 10 SNP GRS in addition to AAb status had the same sensitivity as adding T1D GRS2 and only slightly decreased the specificity (Table 3). This and the possibility of obtaining 10 SNP GRS results in a short time after sampling is another argument for starting to use this parameter routinely, at least for prioritisation for genetic testing. The diagnostic potential of T1D GRS in selecting appropriate candidates for genetic testing remains to be tested in combination with various clinical diagnostic approaches (clinical diagnostic criteria for MODY, the MODY calculator, etc).

### Strengths

We used the GRS in diagnostic process in relatively large homogenous cohort of children, and we also tested two types of GRS in a combination with autoantibodies.

### Limitations

Given the high number of cases and high costs associated with panel sequencing, AAb– patients with high T1D GRS and clinically defined type 1 diabetes were not tested by the panel. Instead, MLPA and Sanger sequencing of the *HNF1A* and *INS* genes were tested only, thus covering thus the most probable genetic causes respective to the patients’ clinical picture.

ZnT8 autoantibodies were tested only in a subset of cases (28 of 71 other AAb negative cases) due to local inaccessibility of this method before year 2018. It is thus possible that some of the AAb– cases would be tested positive for ZnT8 autoantibodies.

The application of T1D GRS2 for identification of monogenic autoimmune subtypes was not investigated in this paper.

The recruitment of patients for the study was not systematic enough to allow estimation of incidence of monogenic diabetes in the studied cohort.

## Conclusions

We have shown that the use of T1D GRS2, in addition to autoantibody testing, is very effective in decreasing the number of unnecessarily genetically tested cases. Children and adolescents with confirmed diabetes and normal BMI, negative autoantibodies and low T1D GRS can be prioritised for genetic testing soon after a diabetes diagnosis, while other cases should be carefully evaluated clinically, taking into consideration AAb levels and other monogenic diabetes criteria due to the natural occurrence of cases with higher GRS2 values and monogenic diabetes.

## Abbreviations

AAb: autoantibody
Aab: autoantibody negative
Aab: autoantibody positive
DKA: diabetic ketoacidosis
GRS: genetic risk score
LADA: Latent autoimmune diabetes in adults
MODY: Maturity Onset Diabetes of the Young
PND: permanent neonatal diabetes
T1D GRS: type 1 diabetes genetic risk score

## Data Availability

Clinical and genotype data can be used to identify individuals and is therefore available only through collaboration to experienced teams working on approved studies.

## Acknowledgments

We appreciate the help of the physicians who performed the clinical examinations and data collection.

## Authors’ contributions

Conceptualization, D.G., J.S.; investigation, Z.D., M.Sk., M.K, M.Sa., J.S, F.G.; writing—original draft preparation, Z.D., M.Sk.; writing—review and editing, M.Sk., J.S., and D.G.; supervision, D.G., M.Sk., J.S.; funding acquisition, M.Sk., J.S.; All authors have read and agreed to the published version of the manuscript.

