## Supplementary Figure 1 for "T1D GRS: a tool for identifying children and adolescents with monogenic diabetes"

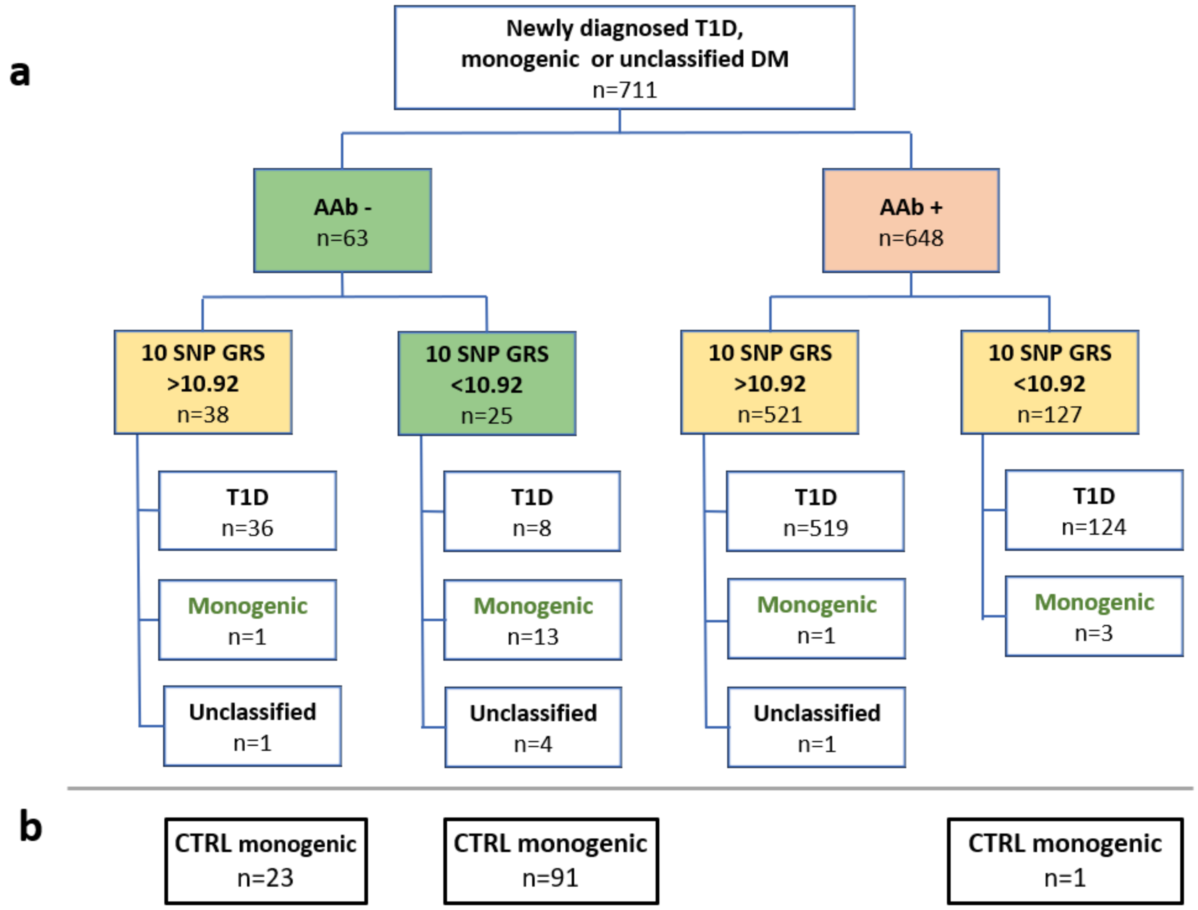


**Supplementary Figure 1:** a) Stratification of newly identified children with diabetes (T2D excluded) according to autoantibodies and 10 SNP GRS; b) Distribution of the monogenic diabetes cases from the control cohort according to autoantibodies and 10 SNP GRS.
